# Epidemiology of SARS-CoV-2 infection in Karnataka State, South India: Transmission dynamics of symptomatic vs. asymptomatic infections

**DOI:** 10.1101/2020.09.17.20196501

**Authors:** Narendra Kumar, Shafeeq K Shahul Hameed, Girdhara R Babu, Manjunatha M Venkataswamy, Prameela Dinesh, Prakash BG Kumar, Daisy A. John, Anita Desai, Vasanthapuram Ravi

## Abstract

**Background:** In this report, we describe the epidemiology of SARS-CoV-2 infection, specifically examining how the symptomatic persons drove the transmission in the state of Karnataka, India, during the lockdown phase.

**Methods:** The study included all the cases reported from March 8 to May 31, 2020 in the state. Any person with history of international or domestic travel from high burden states, those presenting with Influenza-like or Severe Acute Respiratory Illness and high-risk contacts of COVID19 cases, who were SARS-CoV-2 RT-PCR positive were included. Detailed analysis based on contact tracing data available from line-list of the state surveillance unit was performed using cluster analysis software package.

**Findings:** Amongst the 3404 COVID-19 positive cases, 3096 (91%) were asymptomatic while 308 (9%) were symptomatic. Majority of the asymptomatic cases were in the age range of 16-50 years while symptomatic cases were between 31-65 years. Most of those affected were males. Cluster analysis of 822 cases indicated that the secondary attack rate, size of the cluster (dispersion) and occurrence of overt clinical illness is significantly higher when the index case in a cluster was symptomatic compared to an asymptomatic.

**Interpretation:** Our findings indicate that both asymptomatic and symptomatic SARS-CoV-2 cases transmit the infection; however, the main driving force behind the spread of infection within the state was significantly higher from symptomatic cases. This has major implications for policies related to testing. Active search for symptomatic cases, subjecting them to testing and treatment should be prioritized for containing the spread of COVID-19.

**Funding:** Intermediate Fellowship, Wellcome Trust-DBT India Alliance to Giridhara R Babu, Grant number: IA/CPHI/14/1/501499.

## Introduction

India reported the first case of coronavirus disease 2019 (COVID-19) on January 30, 2020, in a medical student who travelled from Wuhan, China^1^. Since then, Severe Acute Respiratory Syndrome Coronavirus 2 (SARS-CoV-2) infection has spread across all states. To date, a total of 5,020,359 SARS-CoV-2 infections have been reported from India. Karnataka, a state in the southern part of India, has a population of 64·41 million spread across 30 administrative units called districts. Karnataka state reported the first case of COVID-19 in an international traveller on March 8, 2020. In the subsequent three months (March 8 to May 31, 2020) the number of COVID-19 cases steadily increased to 3404 in the state. The health authorities of the state initiated laboratory-based surveillance, traditional infection control and public health strategies to contain the spread^2,3^. These included tracing, testing and tracking of contacts and isolating COVID-19 positive subjects^4^. The state implemented several public health measures, including retrospective contact tracing, survey for influenza-like illness and strengthening the laboratory network under the Integrated Disease Surveillance Programme (IDSP). Also, the state had more than 17 task forces set up under the Karnataka State Disaster Management^5^. Several innovative measures were taken to tackle COVID-19 in the initial months. For example, the state started using information from the cell phone tower location from telecom providers and available apps to notify people about their possible interaction with COVID-19 positive persons.

From March 24 2020, the country was under complete lockdown until May 3 2020. On May 21, 2020, India started lifting of lockdown, after which the inter-district travel was permitted within the state of Karnataka. Further, the 3^rd^ lockdown was completely lifted from May 18, 2020, permitting interstate travel. Consequently, there was a huge surge of COVID-19 positive cases from 4^th^ to 27^th^ May by which time most of the residents of Karnataka from other states, especially from those with a high burden of COVID-19 cases returned home. To date of the 3,915,303 samples that have been tested in the state, 475,265 are tested positive (12·14%).

From the beginning of the pandemic, there has been considerable debate on the role of asymptomatic and symptomatic persons in spreading the SARC-CoV-2 infection. Initially, the World Health Organization (WHO) inferred that the role of asymptomatic persons in transmission was minimal^6,7^. However, the WHO acknowledged later that there is a growing body of evidence that even asymptomatic persons can spread the disease^8^. Previous reports suggest that nearly 4-32% of the infections are spread through asymptomatic persons^9,10^. Determining the role of asymptomatic persons in viral transmission in low and middle-income countries is very important. The urban areas in these countries have high population density, poor personal hygiene with ubiquitous crowding.

In this report, we aim to describe the epidemiology of SARS-CoV-2 infection, specifically examining how the symptomatic and asymptomatic persons drive the spread of infection in the state of Karnataka, India, in the lockdown phase.

## Materials and Methods

### Study population

The study included all diagnosed with COVID-19 in the Karnataka state, reported from March 8 to May 31, 2020. Case identification was carried out by the staff of each district, which included the district surveillance officer and their teams, Integrated Disease Surveillance Programme (IDSP) team and the urban health authorities of the major cities. Reporting of Influenza-Like Illness(ILI) and Severe Acute Respiratory Illness (SARI) cases was mandatory for even private health care providers. The contacts of COVID-19 positive people who were in quarantine were monitored.

### Case Definitions

A suspect case of COVID-19 was essentially based on the criteria defined by the Ministry of Health and Family Welfare, Government of India described elsewhere^11^. Briefly, any person with a recent history of international travel (14 days), domestic travel from high burden states of Maharashtra, Gujarat, Delhi and Tamil Nadu and anyone within the state with symptoms of ILI and SARI as well as known high-risk contacts of a confirmed COVID-19 patient were included.

### Laboratory Testing

Testing for SARS-CoV-2 was done initially at five laboratories in the state. This was quickly ramped up to 63 laboratories across the state by May 31, 2020, resulting in an enhanced capacity for testing and identification of COVID-19 cases across Karnataka. Nasopharyngeal and oropharyngeal swabs collected into virus transport medium from suspected cases were subjected to an RT-PCR for detection of SARS-CoV-2 RNA using the Indian Council of Medical Research (ICMR) guidelines for testing^12^. All positive and negative results were entered immediately upon their availability into the ICMR portal and shared with the district/state surveillance teams to facilitate immediate tracing of contacts.

### Data collection

All the sociodemographic, clinical and risk factor details (travel history, symptoms, comorbidities etc.) of patients for every suspected case was collected and entered into a standardized line list by the surveillance teams in the respective districts/cities. The line list also included information about the type of sample, date of sample collection, date of testing, type of RT PCR kit used as well as the test results. All the data was maintained in a centralized database by the State Surveillance Officer. All the data were de-identified before extraction and analysis. A subject who was detected SARS CoV-2 positive was given a P number. The lesser the P number, the earlier the patient was detected either as a source or a contact of a known source.

### Data Analysis

The data analysis from the line list was based on the date of collection of a sample from March 5 to May 31 2020. The trends in positivity rate over time were described by calculating the seven-day moving average. The number tested per million was computed by using State-specific population denominator. The number of individual tests and contacts tested per confirmed case was calculated. Laboratory accessibility was examined by subtracting the date of specimen collection from the specimen receipt date. An indicator for contacts tested per case was estimated by dividing the number of tests among contacts of cases with the number of positive cases. Contact tracing information available in the line list was analyzed to determine the source of infection. Based on the contact tracing information, Cluster Network analysis was performed and presented using Microsoft Excel and Gephi network analysis software^13^. Secondary attack rates were calculated using standard methods and dispersion indices calculated for asymptomatic and symptomatic subjects and expressed as the K factor, which is calculated as variance to mean ratio. This is computed independently of the mean and is robust to changes over time. This index has the potential to be useful in the evaluation of specific health strategies designed to reduce intraregional geographical inequality in the distribution of health status indicators in a specific period^14^.

### Descriptive epidemiology of cases by time, place and person

The frequencies of characteristics of cases were described by age, gender, residence, type of exposure (contact or travel) and symptoms. The presence of any symptoms at the time of specimen collection was also recorded. The date of specimen collection was used to draw the epidemic curve. The time trends were annotated with that of implementation of various public health measures or key events related to the epidemic. (Figure 1).

**Figure 1:**
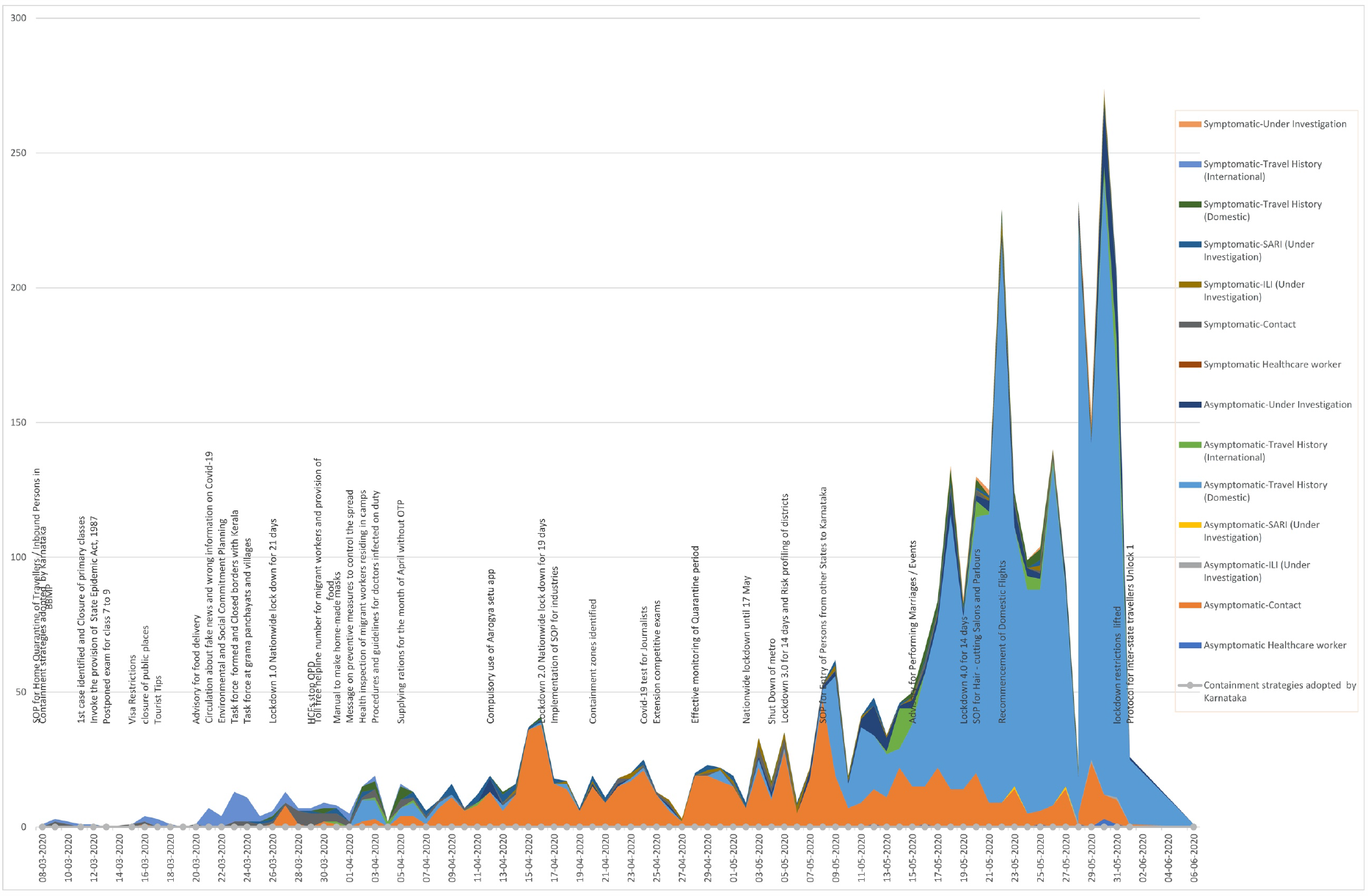
Distribution of case categories, number of cases and interventions undertaken. X axis depicts the timeline while Y axis depicts the number of COVID-19 positive cases. The date wise interventions undertaken by the state of Karnataka is described. The category of patients and the progression in numbers is depicted in various colors in a wave pattern.

### Statistical analysis

All the data were entered into the excel sheet and analyzed using the Statistical Package for Social Sciences (SPSS, version 26) to determine the significance of various parameters. The cluster network analysis was performed using Gephi (version 0·9·2) for analyzing the transmission dynamics. Statistical significance was expressed as a p-value, calculated using chi-square test, t-test and Fischer exact test. Dispersion index was calculated as variance to mean ratio and expressed as K value.

## Results

### Testing

Figure S1 presents the district-wise distribution of cases. The highest number of COVID-19 cases were identified in the capital city of Bengaluru, followed by Kalburgi, Mandya, Yadgiri, Udupi and Raichur. During the study period, the Karnataka state was testing approximately 4377 per million, with a positivity rate of 1·1%. The total cases per million were 2708 in Karnataka. Karnataka followed contact tracing diligently with the highest number of contacts (47) traced for each positive case detected^2^.

### Description of cases by time, place and person

The sociodemographic and salient clinical details of all the COVID-19 cases in Karnataka is presented in Table 1. To understand the dynamics of SARS-CoV-2 transmission within the state, we undertook a detailed analysis of the data available in the line list. Amongst the 3404 COVID-19 positive cases, 3096 (91%) were asymptomatic while 308 (9%) were symptomatic. The age distribution revealed that 54% of the asymptomatic cases were less than 30 years of age compared to 25% of symptomatics in this age group (p<0.00001). On the other hand, 43·5% of the symptomatics were over 46 years of age compared to 18·8% of the asymptomatics in this age group (p<0·00001). The percentage of symptomatics and asymptomatics in the age range 31-45 were more or less similar (31·2% and 29·6% respectively). The gender distribution revealed a male preponderance in both asymptomatic and symptomatic cases.

**Table 1:**
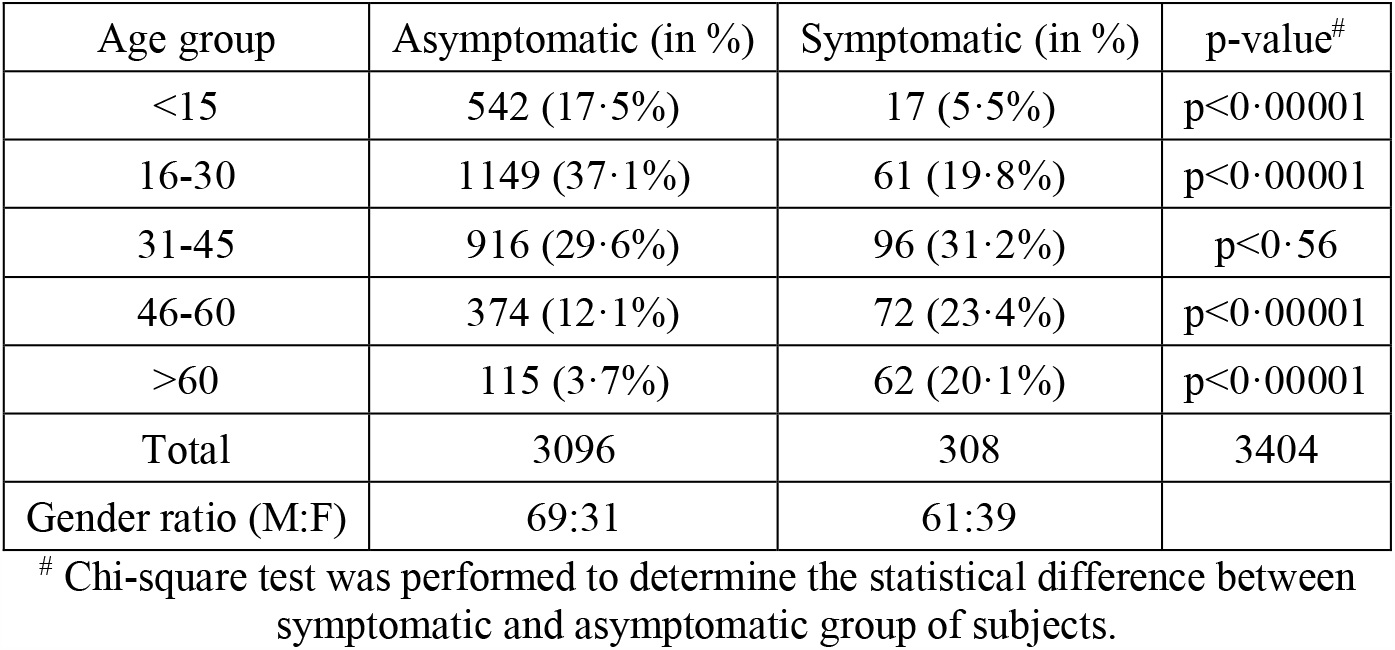
Age group and gender of COVID-19 cases (Mean age 31.26)(N=3404)

Table 2 presents the clinical status and outcome of the COVID-19 cases in Karnataka. Overall, amongst the 3404 cases, 158 (5%) had comorbidities such as diabetes, hypertension, chronic renal disease, malignancy, immunocompromised states etc. Further, the occurrence of comorbidities was ten-fold higher in symptomatic cases (87/308: 28·2%) as compared to asymptomatic cases (71/3096: 2·29%) The overall case fatality recorded in Karnataka was 1.6% (54/3404) with symptomatic cases accounting for 87% (47/54). On the other hand, the mortality amongst asymptomatic cases was 13% (7/54). Overall, 39% of cases (1340/3404) recovered and were discharged from the hospital, while 59% (2010/3404) were active cases as on June 1 2020. (not in table).

**Table 2:**
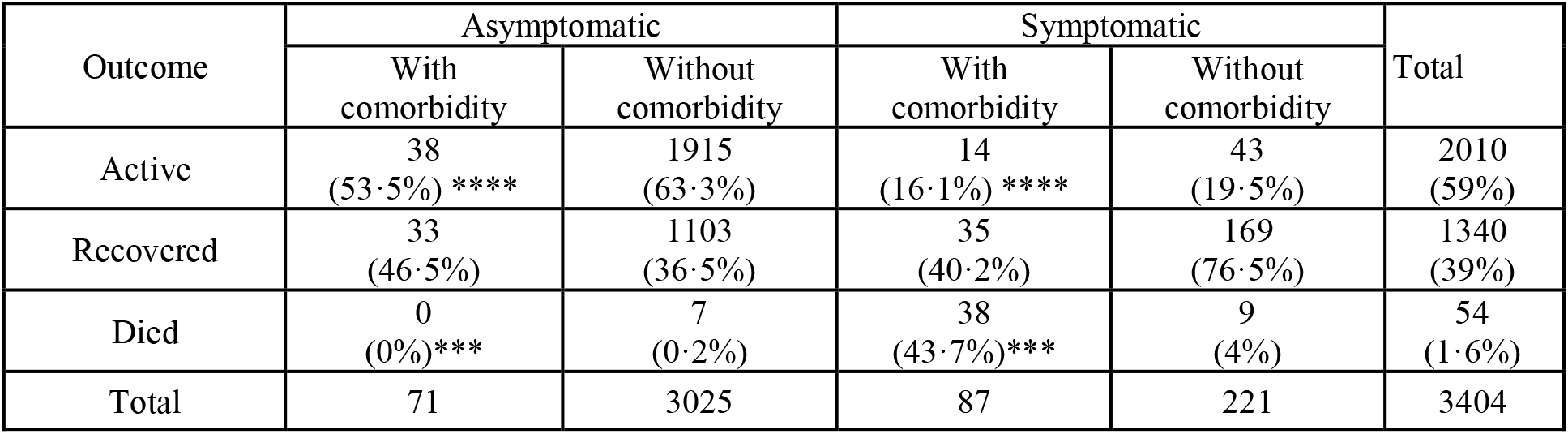
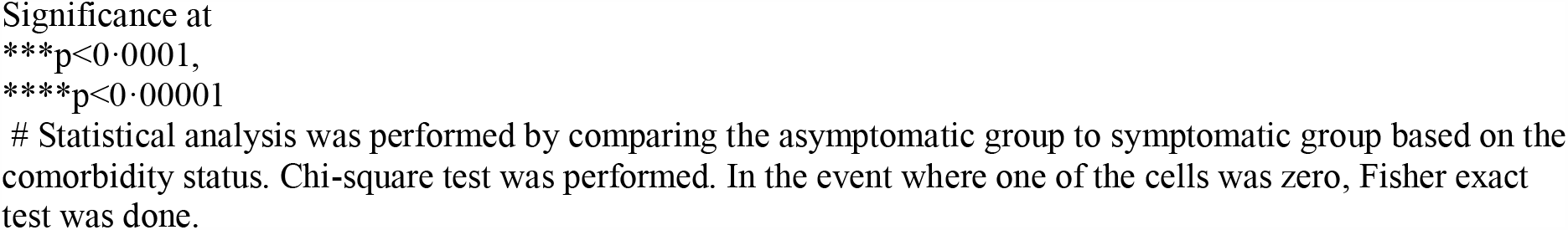
Comorbidity and outcome in COVID19 cases

Among asymptomatic cases with comorbidity, 53·5% were active cases, 46·5% recovered and were discharged while no fatalities were noted. Among symptomatic cases with comorbidity, 16·1% were active cases, 40·2% recovered and were discharged while 43·7% succumbed. Further among asymptomatic cases without comorbidity, the majority (63·3%) were active cases, whereas, among symptomatic cases without comorbidity, the majority (76·5%) recovered and were discharged (Table 2)

Figure 3 shows the distribution of cases for the source in 3137/3404 cases. The acquisition of SARS-CoV-2 infection could mostly be attributed to travel (72·1%); domestic travel (2136, 68%) or international (128, 4%) or as a known contact with a COVID-19 positive case (873, 27·8%). In the remaining 267 cases, the precise reason behind the acquisition of infection could not be ascertained. Amongst these 267 cases (8·3%), 111 presented with ILI / SARI (41·6%) with no information of contact with a COVID-19 positive case. The remaining 156 (58·4%) the source of infection was not traceable. The test positivity of Karnataka mapped against the number of COVID-19 positive cases is depicted in Figure 3. The positivity ranged from 2·7 in March to 0·7 in Mid-April. Subsequently, the positivity rate was 2·1. (Figure3)

**Figure 2:**
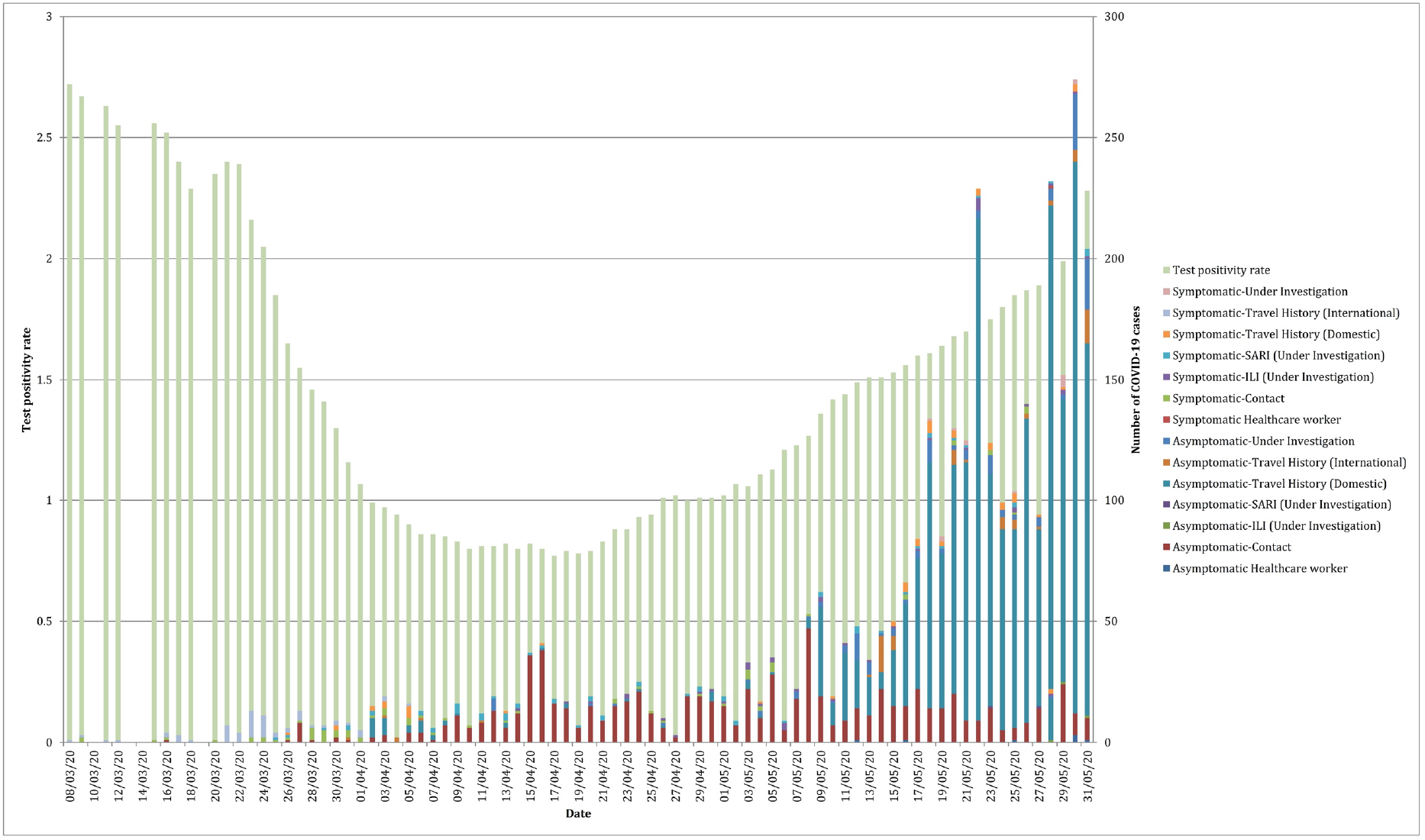
Distribution of case categories and test positivity. X axis depicts the timeline, Y axis depicts the test positivity rate while the Z axis depicts the number of COVID-19 positive cases. Note there was a huge surge in the number of COVID-19 positive cases in the second half of May 2020.

**Figure 3:**
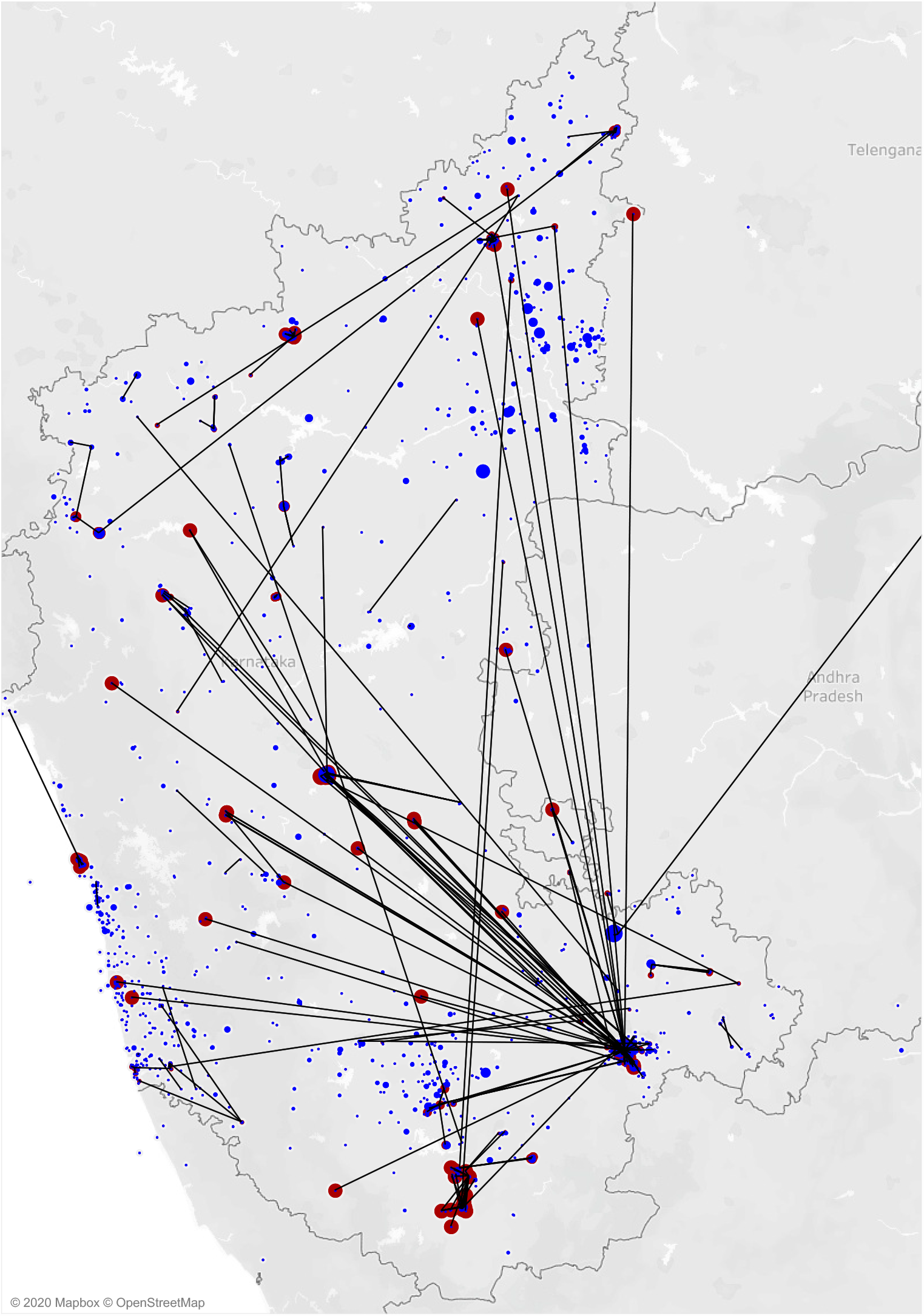
Distribution and direction of transmission of COVID-19 cases in Karnataka State. Each dot represents a person with COVID-19. Primary cases are in red and secondary cases in blue. Blue coloured dots connected to primary cases using lines. The dots not connected by lines represent independent introduction from elsewhere. Bengaluru city was the epicentre, had the highest density of COVID-19 cases, from where spread occurred to several districts in the state.

As evident from Figure 3, the cases started from Bengaluru and have transmitted to people in the other districts, either by contact in Bengaluru or by travelling to other districts The other hotspot districts were Gulbarga, Mysore and Dakshina Kannada, which had an independent source of infection and local spread. The majority of the COVID-19 cases (68·5%; 2264/3304) had a history of either international (5·7%; 128/2264) or domestic travel (94·3% 2136/2264) which implies that this was the main mode of introduction of SARS-CoV-2 into the state.

Amongst the 873 cases with a history of contact with a COVID-19 positive case, 822 could be epidemiologically linked to 144 source cases. The remaining 51/873 had a history of contact but could not be precisely linked to a known source. A network cluster analysis of 46 index cases identified as a source along with the 659 secondary cases is presented in Table 3 and Figure 4. As evident from Table 3, the size of each cluster ranged from 5-58 cases per cluster and in the majority of the large clusters (>5 cases), the source patient was a symptomatic case (Figure 4). The proportion of secondary cases manifesting overt clinical illness was higher when the index case was symptomatic. Amongst the 545 secondary cases, 43 were symptomatic, and 616 were asymptomatic. The 43 symptomatic cases resulted in the transmission of infection to 659 cases giving a mean secondary attack rate of 17·03 (range 5-56). On the other hand, the 616 asymptomatic source cases contributed to a total of 504 secondary infections giving a mean attack rate of 3·01 (range 5-22). Similarly, the index of dispersion also was significantly higher when the source case was symptomatic (14·03) as compared to the source case being asymptomatic (3·03). However, the number of cycles of transmission were not statistically different for symptomatic and asymptomatic cases (mean number of cycles).

**Table 3.** Cluster analysis depicting the source of infection, secondary attack rate and dispersion of COVID19 cases.

| Index case status | Total no. of Clusters (n=46) | Total no. of subjects associated with the cluster (n=659) | Clinical status of secondary cases in clusters (n=659) ^ |  | Average no. of subjects per cluster (Range)<sup>+</sup> | Index of dispersion (K value) |
| --- | --- | --- | --- | --- | --- | --- |
|  |  |  | Symptomatic (n=43) | Asymptomatic (n=616) |  |  |
| Symptomatic | 32 (69%) | 545 (82.7%) | 41 | 504 | 17.03 (5-56) | 14.3 |
| Asymptomatic | 14 (31%) | 114 (17.3%) | 2 | 112 | 8.1 (5-22) | 3.03 |

Statistical significance between symptomatic and asymptomatic index cases using Chi-square test and t-test for dependent means and ^ $p=0.02$ (chi-square test) + $p=0.00005$ (t-test)

**Figure 4:**
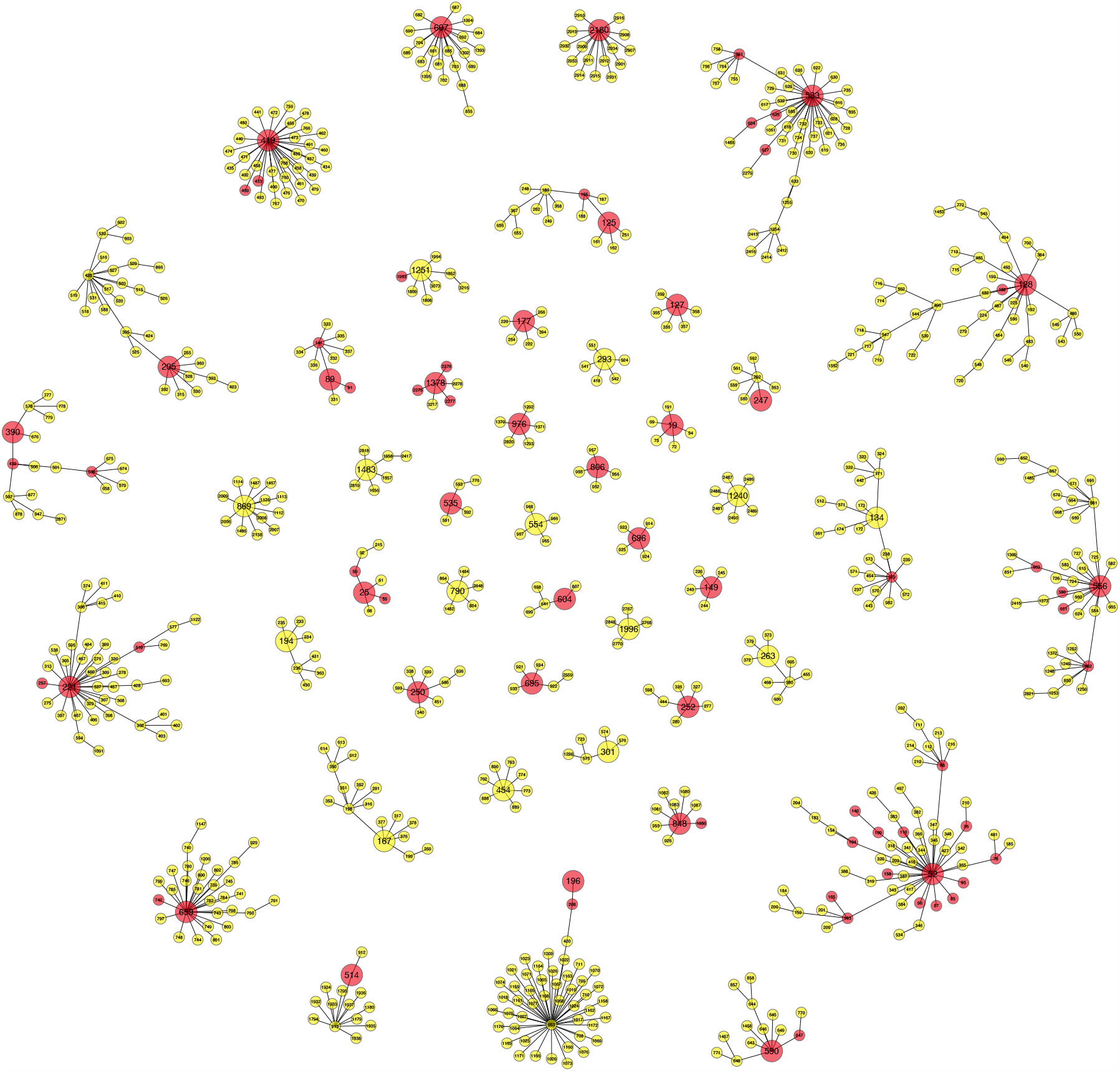
Cluster diagram depicting symptomatic and asymptomatic COVID-19 cases in the Karnataka state, March-May 2020. Each case is represented as a dot: red dot indicates a symptomatic case while yellow dots represent an asymptomatic case. The centre of each cluster is a source case indicated with the P number of the index case. The lesser the P number, the earlier the patient was detected either as a source or a contact of a known source. Note there were 46 clusters with five or more cases. The index case of majority (32/47) of clusters was a symptomatic patient (red). The number of cycles of transmission was higher (Mean 2·2) when the index was a symptomatic case compared to an asymptomatic (mean 1·6).

## Discussion

This study describes the epidemiology of SARS CoV2 infections in the state of Karnataka, South India. Before March 8 2020, there were no COVID-19 cases in Karnataka. SARS CoV-2 infection was introduced into the state by people travelling from foreign countries until the country went into lockdown in the fourth week of March 2020 when international flights were not allowed into the country. What initially started as an infection in the capital city of Bengaluru at the beginning of March 2020, quickly spread to the other parts of the state. The majority of cases were mainly (68.5%; 2264/3304) spread due to domestic (94·3%, 2136/2264) and international travel (5·7%; 128/2264). One of the major reasons attributable to the spread of the infection across the state was domestic travellers arriving into Karnataka just before the lockdown from other states of India (Figure 1). Indeed, the majority of the COVID-19 cases had a history of either international or domestic travel, which implies that this was the main mode of introduction of SARS-CoV-2 into the state.

From the detailed epidemiological analysis of data in this study, it emerges that both symptomatic and asymptomatic cases contributed to the transmission and spread of SARS-CoV-2 infection within the state. An overwhelming majority of the COVID-19 cases in Karnataka were asymptomatic at the time of specimen collection, and this is similar to the results reported in India as well as other Asian countries^15^ and in stark contrast to what is reported in several western countries^16^. Most of the asymptomatic cases were young adults or middle-aged (<50 years of age −20; Median, interquartile range), with a very small percentage having comorbidities. Consequently, the mortality rate was low, and the recovery rate was very high amongst this group (Table 2). Using this core information, we carried out a detailed analysis of our data to calculate secondary attack rates and perform network cluster analysis to understand the dynamics of SARS-CoV-2 transmission within the state.

Our analysis suggests that symptomatic cases were the prime drivers of the SARS-CoV-2 transmission within the state. The evidence for this is manifold; firstly the mean secondary attack rate was 7·1 for symptomatic cases (90 symptomatic source cases transmitted infection to 645) as compared to 3·1 for asymptomatic cases (54 asymptomatic source cases transmitted infection to 177), and this difference was statistically significant. Secondly, the dispersion index for symptomatic source cases was fourfold higher (14·3) as compared to asymptomatic source cases (3·03). Thirdly, network cluster analysis (Figure 3) revealed that symptomatic source patients contributed to 33 large clusters (>5 cases) with the mean size of each cluster being 17·03 (range 5-56 cases per cluster). In contrast, the asymptomatic source cases contributed to 14 large clusters (>5 cases) with the mean size of each cluster being 8·1 (range 5-22 cases per cluster). Further, the proportion of secondary cases in the clusters manifesting overt clinical illness was higher when the index case was symptomatic compared to asymptomatic. Also, the mortality was higher among the symptomatic, especially among the aged and with comorbidities. Therefore, our results indicate that public health actions focused on testing, tracing, tracking and treating the symptomatic person must be accorded top priority in curbing the transmission and in reducing the mortality.

There were two limitations to this study. A key factor in the transmission of COVID-19 is the high level of shedding of SARS-CoV-2 virus from the upper respiratory tract. Viral RNA shedding is higher at the time of symptom onset and declines after days or weeks^17^. The difficulty of distinguishing asymptomatic cases from those who are pre-symptomatic is a major stumbling block. Since longitudinal data on the occurrence of symptoms after the date of collection of specimens was not available in this study, the proportion of asymptomatic cases in this study may indeed be lower than 90%. Complete contact tracing was available for cluster network analysis only in 966/3404 (34%) COVID-19 cases until May 31 2020.

In conclusion, the findings of this study have major implications for policies related to testing and treatment. Active search for symptomatic cases, subjecting them to testing and treatment will be key to containing the spread of COVID-19 in the community. Above all, public health messages should emphasize on the higher chances of acquiring SARS CoV-2 infection from a symptomatic individual as well as the potential of symptomatic cases to transmit the infection to a larger number of immediate contacts.

## Data Availability

Not Applicable

## Acknowledgements

The authors acknowledge the COVID laboratory and data management team of Dept. of Neurovirology, NIMHANS (in alphabetical order): John Bannerjee, Tina Damodar, Priti Das, Anson George, Kiran Hosallimath, Ruthu Nagaraju, Tanmoy Nandi, Amrita Pattanaik, Chitra Pattabhiraman, Harsha PK, Risha Rasheed, Vijaykiran Reddy, Vijayalaksmi Reddy, Reeta S Mani, Ashwini Shetty, Sourabh Suran as well as all the staff of the ICMR approved SARS-CoV-2 testing laboratories in the state of Karnataka. The contributions of the state and district surveillance officers and their teams are also gratefully acknowledged.

## Author contributions

NK and SKSH contributed equally to data collection, analysis and preparation of manuscript, GRB contributed to analysis and review of manuscript, MMV contributed to laboratory testing, PD and PKBG contributed to collection, collation of data and preparation of line-list, DAJ contributed to data analysis and figures, AD contributed to data analysis, preparation and review of the manuscript and VR conceptualized the study and contributed to data analysis, preparation and review of the manuscript.

## Declaration from the authors

GRB and DAJ are employees of Indian Institute of Public Health, Bengaluru, Public Health Foundation of India and their employer had no say in the design of the study or the decision to publish. All other authors are employees of either state government (PD, PKBG) or NIMHANS (Central government). The employers had no role in the design of the study or the decision to publish. The authors declare that they do not have any other financial or non-financial relationships that could present a conflict of interest.

